# Long-term Public Healthcare Burden Associated with Intimate Partner Violence among Canadian Women: A Cohort Study

**DOI:** 10.1101/2024.06.18.24309101

**Authors:** Gabriel John Dusing, Beverley M. Essue, Patricia O’Campo, Nicholas Metheny

## Abstract

Intimate partner violence (IPV) is a major global health issue, yet few studies explore its long-term public healthcare burden in countries with universal healthcare systems. This study analyzes this burden among Canadian women using data from the Neighborhood Effects on Health and Wellbeing survey and Ontario Health Insurance Plan (OHIP) records from 2009-2020. We employed inverse probability weighting with regression adjustment to estimate differences in cumulative costs and OHIP billings between those reporting exposure to IPV during the survey and those who did not. Our sample included 1,094 women, with 38.12% reporting IPV exposure via the Hurt, Insult, Threaten, Scream scale. Findings show a significant public healthcare burden due to IPV: women reporting IPV in 2009 had an average of 17% higher healthcare costs and 41 additional OHIP billings (0.1732;95% CI: 0.0578-0.2886; 41.23;95% CI: 12.63-69.82). Policies prioritizing primary prevention and integration of trauma-informed care among healthcare providers are vital to alleviate the long-term burden on public health systems.

## Background

Upwards of 40% of cisgender, Canadian women have experienced some form of intimate partner violence (IPV) in their lifetime (1,2). Including aggressive, coercive, controlling, or abusive behaviors resulting in physical and non-physical harm (3), IPV occurs across social strata (4,5) and outside male-female relationships (6–8). However, the most widespread form of IPV is violence perpetrated against cisgender women by a cisgender male intimate partner (2,9).

IPV exposure is linked to health effects through causal pathways involving physical and psychological trauma, chronic stress, and control (Figure 1) (10). First, IPV may cause physical and psychological trauma, directly increasing the risk of physical disability (11) and poor mental health (9,12–17). Long-term, chronic stress may trigger endocrine and immune responses, leading to inflammation (18–20) and non-communicable diseases (6,11,21–24). Restrictions on autonomy are shown to lead to untreated sexually transmitted infections and gynecological issues (13,25,26) and delaying disclosure of health issues, impeding treatment of health conditions (27,28).

**Figure 1:**
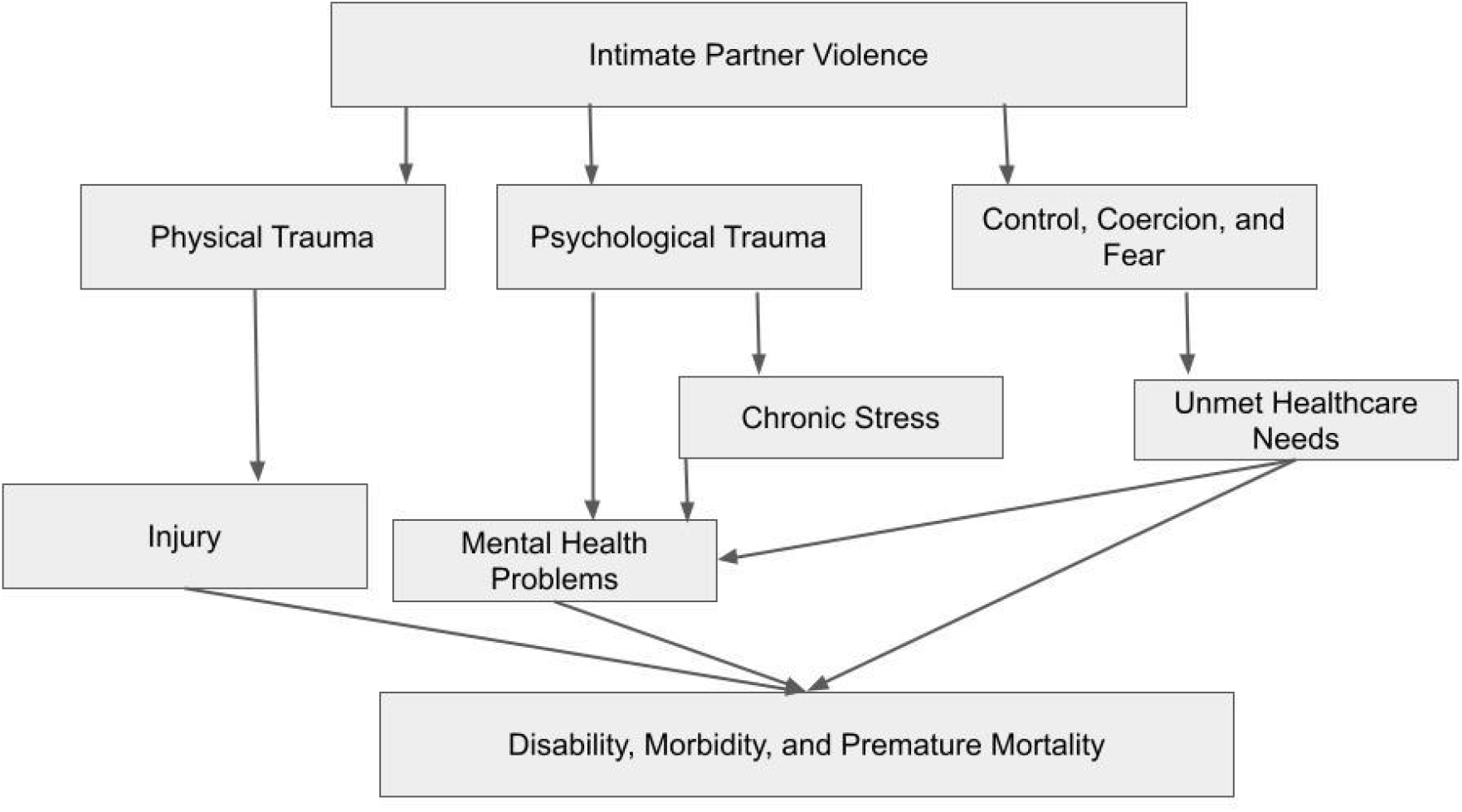
Theoretical Mechanism Linking IPV Exposure to Negative Long-term Health Trajectories

Empirical evidence supports the association between IPV exposure and long-term negative health effects. A 2021 study linking Manitoba court records with healthcare data showed that women exposed to IPV had a nearly sevenfold greater risk of physical injuries, with incidences of 55.8 per 1000 (exposed) versus 8.5 per 1000 (non-exposed) (29). A longitudinal Australian study showed that women exposed to IPV had consistently poorer general health over 16 years compared to non-exposed women (30). Long-term societal costs associated with spouse-perpetrated IPV were estimated to be CAD$7.4 billion in 2009 (31), which is likely an underestimate given the underreporting of IPV (32–34), the limited scope of the study, and data constraints. Although some cost estimates of IPV exposure in countries with universal healthcare exist (35–37), extant literature centers the United States, limiting generalize to jurisdictions that have universal healthcare systems. Furthermore, much of the existing research in universal healthcare systems are cross-sectional (11,27,28,38), rely on a single item to identify IPV (28,35,36), use administrative records to capture the most severe forms of IPV (29,37,39), or is limited by small sample sizes (19,28).

This study aims to compare differences in cumulative cost and total number of billings by IPV exposure over ten years among a representative sample of women residing in Toronto, the most populous and diverse city in Canada (40). We hypothesize women who report IPV exposure will have greater public healthcare costs and more frequent billings over the subsequent ten years compared to women who did not report IPV in this context.

## Methods

### Study Cohort

The study sample was derived from the Neighbourhood Effects on Health and Well-being (NEHW), a cross-sectional study of 2,412 English-speaking residents (age 25-64) from 50 Toronto neighborhoods collected over 2009-2011 (41). NEHW had high response (>80%) and completion (>96%) rates, and utilized multistage random sampling to ensure the sample was representative of the socio-demographic and economic profile of Toronto. Participants from the NEHW study eligible for our analysis were (1) female; (2) completed the Hurt, Insult, Threaten, Scream (HITS) screener for IPV; and (3) provided a valid Ontario Health Insurance Plan (OHIP) number at the time of survey completion. We then contracted ICES, the organization authorized by the Ontario government to house and link provincial health administrative data (42,43), to link these participants to their personal OHIP records from the time of survey completion (2009–2011) to the latest year available (2020). OHIP is Ontario’s provincial healthcare plan, providing universal coverage for medical expenses, including all physician, emergency department, and hospital visits (44). All Canadian citizens, permanent residents, and non-citizens with employment authorization aged 25+ residing in Ontario are eligible for OHIP (45). The final study sample consisted of N=1,094 participants.

### Exposure

NEHW queried participants about their experiences with IPV over the past decade using the HITS scale (46–48). HITS has high sensitivity (96%) and specificity (91%) (47) and evaluates IPV through four key behaviors: physical aggression, verbal insults/degradation, threatening harm, and yelling. Participants rate the occurrence of each behavior using a five-point Likert ranging from 1 - "never" to 5 - "frequently." The summed scores (range: 4-20) identify the presence and frequency of IPV, with a cutoff of 10 indicating a threshold of IPV exposure (47). Individuals meeting this cutoff formed the exposure group, while those who did not were designated as the comparison group.

### Outcome

The main outcome of this study is the public healthcare burden assessed through the 1) cumulative costs; and 2) cumulative number of billings to OHIP, First, each survey participant’s healthcare costs were measured through the natural log of each participant’s cumulative OHIP expenditure (adjusted to 2019 values) from the time of survey completion to the end of the study period (March 31, 2020). The natural log of cumulative cost allows interpretion as a percentage change, facilitating comparability with other studies. Second, we summed the cumulative number of healthcare billings to OHIP over the study period for each participant.

### Covariates

Statistical models used to adjust regression models (main analyses) and calculate propensity scores (supplementary analyses) included the following demographic and socio-economic characteristics known to be correlated with IPV exposure: year of birth (continuous); length of OHIP eligibility (continuous); has children (yes/no); and total household income in Canadian dollars (<$15,000, $15,000 - $29,999, $30,000-$49,999, ≥ $50,000, or not reported). Immigrant status (born in Canada or not) was included for being correlated with differences in health status (49,50) as was healthcare utilization (49,51,52). The Charlson Comorbidity Index (CCI) was included to capture the burden of chronic and comorbid conditions and was computed using OHIP records (53–55). The Ontario Marginalization Index (ON-Marg) of neighborhood-level material deprivation (continuous) (56), encapsulate difficulties in meeting essential needs and is calculated via metrics including income, housing quality, and educational attainment; such measures are linked to differential patterns of healthcare utilization (46,57) and IPV risk (58–60).

### Analyses

Descriptive analyses assessed the association between sample characteristics and IPV exposure, using chi-square tests for categorical variables, t-tests for differences in means, and bivariate quantile regression for differences in medians (Table 1). We also included a table with the distribution of the top ten most common billing codes categorized by IPV exposure at survey completion (Table 2), highlighting where the health system’s IPV costs are aggregated. Across all analyses, statistical significance was set at alpha=0.05 and conducted using Stata 17 (61).

**Table 1:** Sample characteristics at time of survey participation, unmatched (N=1,094)

|  | No IPV Reported within 10-years of Survey Participation |  | Screened Positive for IPV within 10-years of Survey Participation |  | Total |  |
| --- | --- | --- | --- | --- | --- | --- |
|  | *N* | *Row %* | *N* | *Row %* | *N* | *Row %* |
| **Totals** | 677 | 61.88 | 417 | 38.12 | 1,094 | 100 |
|  | *Median* | *Interquartile Range* | *Median* | *Interquartile Range* | *Median* | *Interquartile Range* |
| **Cumulative Healthcare Cost (Standardized to 2019 Inflation)** | 34,997 | [19001,64320] | 37,839 | [21873,74140] | 35,745 | [20072,67481] |
| **Cumulative OHIP Billings** | 218 | [127,346] | 221 | [140,353] | 219 | [133,351] |
| **Neighborhood Material Deprivation Index** | -0.24 | [-0.73,0.55] | 0.02 | [-0.58,0.86] | -0.11 | [-0.70,0.70] |
|  | *Mean* | *Standard Deviation* | *Mean* | *Standard Deviation* | *Mean* | *Standard Deviation* |
| **Log Total Years of OHIP Eligibility** | 12.41 | 1.37 | 12.44 | 1.30 | 12.42 | 1.35 |

|  | *N* | *Column %* | *N* | *Column %* | *N* | *Column %* |
| --- | --- | --- | --- | --- | --- | --- |
| **Age** |  |  |  |  |  |  |
| 25-29 | 33 | 4.87 | 34 | 8.15 | 67 | 6.12 |
| 30-39 | 94 | 13.88 | 94 | 22.54 | 188 | 17.18 |
| 40-49 | 180 | 26.59 | 124 | 29.74 | 304 | 27.79 |
| 50-59 | 224 | 33.09 | 120 | 28.78 | 344 | 31.44 |
| 60+ | 146 | 21.57 | 45 | 10.79 | 191 | 17.46 |
| **Marital Status** |  |  |  |  |  |  |
| Unmarried | 271 | 40.03 | 166 | 39.81 | 437 | 39.95 |
| Married | 406 | 59.97 | 251 | 60.19 | 657 | 60.05 |
| **Has Children** |  |  |  |  |  |  |
| No | 247 | 36.48 | 121 | 29.02 | 368 | 33.64 |
| Yes | 430 | 63.52 | 296 | 70.98 | 726 | 66.36 |
| **Born in Canada?** |  |  |  |  |  |  |
| Yes | 416 | 61.45 | 281 | 67.39 | 697 | 63.71 |
| No | 261 | 38.55 | 136 | 32.61 | 397 | 36.29 |
| **Employment Status** |  |  |  |  |  |  |
| Unemployed | 202 | 29.84 | 120 | 28.78 | 322 | 29.43 |
| Employed | 475 | 70.16 | 297 | 71.22 | 772 | 70.57 |
| **Total Household Income (2009 CAD)** |  |  |  |  |  |  |
| <15,000 | 15 | 2.22 | 14 | 3.36 | 29 | 2.65 |
| 15,000 - 29,999 | 40 | 5.91 | 27 | 6.47 | 67 | 6.12 |
| 30,000-49,999 | 79 | 11.67 | 55 | 13.19 | 134 | 12.25 |
| ≥ 50,000 | 501 | 74.00 | 307 | 73.62 | 808 | 73.86 |
| Not reported | 42 | 6.20 | 14 | 3.36 | 56 | 5.12 |
| **Charlson Comorbidity Index** |  |  |  |  |  |  |
| 0 | 125 | 18.46 | 116 | 27.82 | 241 | 22.03 |
| 1-2 | 333 | 49.19 | 208 | 49.88 | 541 | 49.45 |
| 3-4 | 132 | 19.50 | 54 | 12.95 | 186 | 17.00 |
| 5+ | 87 | 12.85 | 39 | 9.35 | 126 | 11.52 |

**Table 2:**
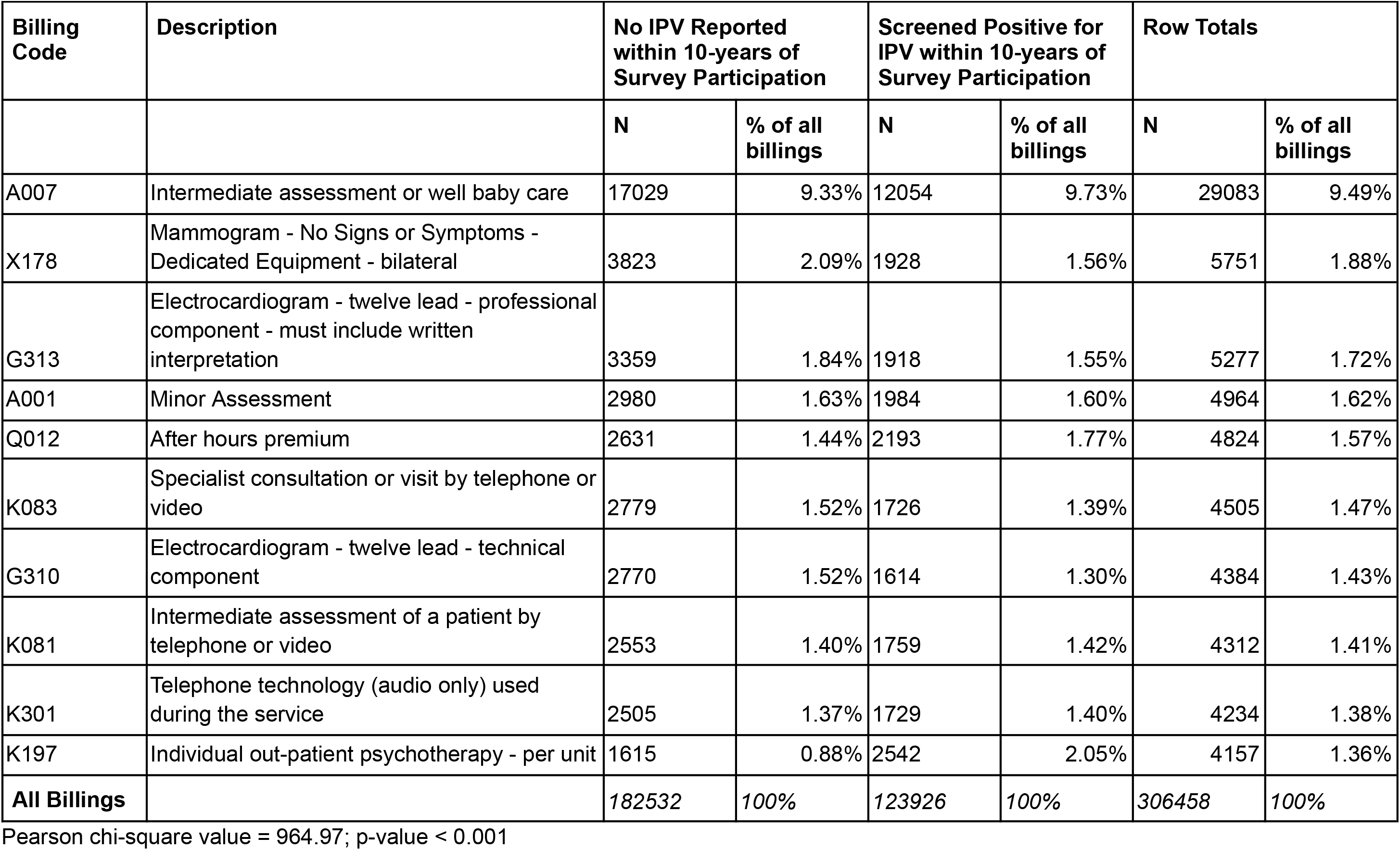
Distribution of the top 10 most frequent OHIP billing codes by IPV status over the study period (2011-2020)

| Billing Code | Description | No IPV Reported within 10-years of Survey Participation |  | Screened Positive for IPV within 10-years of Survey Participation |  | Row Totals |  |
| --- | --- | --- | --- | --- | --- | --- | --- |
|  |  | N | % of all billings | N | % of all billings | N | % of all billings |
| A007 | Intermediate assessment or well baby care | 17029 | 9.33% | 12054 | 9.73% | 29083 | 9.49% |
| X178 | Mammogram - No Signs or Symptoms - Dedicated Equipment - bilateral | 3823 | 2.09% | 1928 | 1.56% | 5751 | 1.88% |
| G313 | Electrocardiogram - twelve lead - professional component - must include written interpretation | 3359 | 1.84% | 1918 | 1.55% | 5277 | 1.72% |
| A001 | Minor Assessment | 2980 | 1.63% | 1984 | 1.60% | 4964 | 1.62% |
| Q012 | After hours premium | 2631 | 1.44% | 2193 | 1.77% | 4824 | 1.57% |
| K083 | Specialist consultation or visit by telephone or video | 2779 | 1.52% | 1726 | 1.39% | 4505 | 1.47% |
| G310 | Electrocardiogram - twelve lead - technical component | 2770 | 1.52% | 1614 | 1.30% | 4384 | 1.43% |
| K081 | Intermediate assessment of a patient by telephone or video | 2553 | 1.40% | 1759 | 1.42% | 4312 | 1.41% |
| K301 | Telephone technology (audio only) used during the service | 2505 | 1.37% | 1729 | 1.40% | 4234 | 1.38% |
| K197 | Individual out-patient psychotherapy - per unit | 1615 | 0.88% | 2542 | 2.05% | 4157 | 1.36% |
| **All Billings** |  | **182532** | **100%** | **123926** | **100%** | **306458** | **100%** |

Pearson chi-square value = 964.97; p-value < 0.001

We used inverse probability weighting with regression adjustment (IPWRA) – a matching method – to assess the association between outcome and exposure. IPWRA has previously been used to evaluate the effect of financial programs in Mexico (62) and Bangladesh (63), and household access to clean water in India (64) on the risk of IPV among women in these countries. Matching with IPWRA has two stages. First, a model for the exposure status is fitted to obtain propensity scores representing the likelihood of treatment assignment, which are then used to weight each individual in the exposure group and their counterpart in the comparison (65). The second stage applies regression adjustment using these weights to estimate average treatment effects (ATE) and average treatment effects on the treated (ATET) to compute averages in the predicted outcomes (i.e., cumulative billings and log cumulative cost) by exposure status (IPV vs no IPV). The ATE measures the expected outcome differences between the exposure and comparison groups across the entire population, regardless of actual exposure status (66,67). The ATET measures expected outcome differences solely within the exposure group. The alignment of these estimates suggests that the findings may generalize beyond the study sample, within matched characteristics (68).

IPWRA is similar to other matching methods in that it balances the IPWRA-weighted distribution of observed covariates between exposure and control groups, approximating a randomized experiment using observational data. However, IPWRA is characterized as “doubly robust” for estimating treatment effects since it requires correct specification of only either the outcome or the propensity score model (69). For each model, covariate balance is assessed through a table of standardized differences and variance ratios for each covariate. Standardized differences between zero and 0.10 (70) and variance ratios close to one indicate negligible differences between exposure/comparison groups and attainment of covariate balance (67,70,71).

### Sensitivity Analyses

To ensure the robustness of our results, conventional propensity score matching was conducted with the same outcomes and covariates as in our main analysis, consistent with methods from prior studies (62,64).

## Results

### Sample Characteristics

Table 1 presents key characteristics of the unmatched study sample by IPV exposure. Of the N=1,094 women in our study sample, 38.12% screened positive for IPV via the HITS instrument within the decade before participating in NEHW. The median cumulative cost to OHIP over the study period for the exposure group was CAD$37,839 (IQR: [21873,74140]), while the median cumulative cost of the comparison group was CAD$34,997 (IQR: [19001,64320]). Likewise, the exposure group had more cumulative OHIP billings (median: 221; IQR: [140,353]) than the comparison group (median: 218; IQR: [127,346]). Although the exposure group appeared to have higher median cumulative OHIP costs and billings than the comparison, these unadjusted differences did not reach statistical significance.

Material deprivation, age, having children, immigrant status, and CCI were all found to be significantly associated with exposure status in univariate analyses, justifying their inclusion in the weighting stage of the IPWRA analysis. Exposed women appeared to live in significantly (p=0.001) more materially deprived neighborhoods indicated by a higher median material deprivation index (median:0.02; IQR: [-0.58,0.86]) than comparison group counterparts (median:-0.24; IQR: [-0.73,0.55]). The exposure group was younger with 30.69% of women under 40 years old, as opposed to only 18.75% in the comparison group (p<0.001). Additionally, 70.98% of the exposure group had children, contrasted with 63.52% of the comparison group (p=0.025). The exposure group (relative to the comparison group) had a higher proportion of those with a CCI of zero (27.82% vs 18.46%) (p<0.0001).

Further contextualizing healthcare billings by IPV exposure, Table 2 presents differences in the distribution of the top ten most common OHIP billings (72,73) by IPV status. Billing distribution appears to differ significantly by exposure (p<0.0001), most notably a higher frequency of billings for K197 “Individual out-patient psychotherapy - per unit.” This code constituted 0.88% of billings in the comparison group and 2.05% of the exposure group.

### Model Results

The IPWRA estimates for ATE and ATET are reported in Table 3. On average across the entire study population, assuming universal IPV exposure in a counterfactual scenario, the ATE was 0.1732 (95% CI: 0.0578-0.2886) for log cumulative cost and 41.23 (95%CI: 12.63-69.82) in cumulative billings, indicating a 17.32% elevation in healthcare costs and 41.23 additional billings over the ten-year study period. Meanwhile, the ATET, that is, the average effect among women in our study who screened positive for IPV using HITS, was 0.1731 (95% CI: 0.0595-0.2866) and 47.90 (95%CI: 18.54-77.27) for log cumulative cost and cumulative healthcare billings respectively. This indicates that women in our study sample who screened positive for IPV had healthcare costs that were approximately 17.31% higher than what they would have been had they not experienced IPV, and 47.90 additional billings. The similarity of the ATE and ATET results for both outcomes suggests that the treatment effect is likely generalizable to women in the wider population (68).

**Table 3:** Estimated Average Treatment Effects (ATE) and Average Treatment Effects on the Treated (ATET) for outcomes over the study period (2011-2020) estimated with Inverse Probability Weighting with Regression Adjustment. Exposure is IPV within 10-years of survey participation.

|  | Estimate | 95% Confidence Interval |  |
| --- | --- | --- | --- |
|  | Log of Cumulative Healthcare Cost |  |  |
| ATE | 0.1732** | 0.0578 | 0.2886 |
| ATET | 0.1731** | 0.0595 | 0.2866 |
|  | Cumulative Number of OHIP Billings |  |  |
| ATE | 41.23** | 12.63 | 69.82 |
| ATET | 47.90** | 18.54 | 77.27 |

\*p-value < 0.05, \*\*p-value < 0.01, \*\*\*p-value < 0.001

Supplementary Table 1 presents the standardized differences and variance ratios between covariates in the exposure and comparison groups, in both the matched and unmatched sample, for all four models: one estimating ATE and ATET for each outcome (cumulative OHIP billings and log cumulative OHIP cost). Covariate distribution was improved after application of IPWRA-derived weights, with most standardized differences falling below the pre-specified cutoff (0.10) (70) and variance ratios close to one.

The results derived from conventional propensity score matching are presented in Supplementary Table 2. The ATE and ATET for both cumulative OHIP billings and log cumulative cost were consistent with our main analyses, albeit with reduced magnitude. Specifically, the ATE and ATET for log cumulative cost indicate 20.42% (95%CI: 7.37%-33.47%) and 19.20% (95%CI: 4.45%-33.95%) higher OHIP costs, respectively. Cumulative OHIP billings indicate an additional 34.87 billings (95%CI: 3.33-66.41) for ATE and 60.00 billings (95%CI: 26.54-93.45) for ATET.

## Discussion

This study is among the first to assess the public healthcare burden associated with IPV-exposure using a representative sample directly linked to data from a universal healthcare provider. The exposure group, i.e., those who screened positive for IPV during the ten years before participating in NEHW, had 17% higher public healthcare costs and 43 additional healthcare billings over the subsequent 10-year period than the matched comparison group. The Canadian Institute for Health Information estimated that age- and sex-adjusted annual OHIP spending was $5,248 per person (74). Based on our analysis, such IPV exposure incurs an additional $892 per affected individual (17% of $5,248). With Ontario’s female population holding steady at around 7 million since 2011 (75), and assuming a conservative 13% have experienced IPV in the past year (76), the excess OHIP costs amount to approximately CAD$811 million annually, or an additional CAD$8.11 billion over the study period.

### Comparison with Prior Literature

A prior American study found women experiencing lifetime IPV had 19% (95%CI: 13%-26%) higher healthcare costs than those who did not (77). Despite similar excess cost estimates, their cohort was derived from enrollees in a single health maintenance scheme, while ours is based on a representative sample linked to healthcare data from a universal provider. This distinction is crucial, where a publicly funded health system in Ontario, offering universal coverage, contrasts with a more selective single health maintenance organization in the US.

We found that the exposure group received outpatient mental healthcare at higher rates than the comparison group (2.05% vs. 0.88%); potentially explained by higher rates of poor mental health outcomes found in prior studies (12,78), for example, exposed women had 1.72 (95%CI: 1.28-2.31) higher odds of subsequent depressive symptoms, and 2.19 (95%CI: 1.39-3.45) higher odds of postpartum depression (14).

### Strengths and Limitations

This study has several major strengths. The average follow-up time was 9.5 years. Follow-up was conducted using OHIP records, minimizing loss to follow-up except in cases of emigration from Ontario or death. Our study sample was drawn from Canada’s largest and most diverse city using rigorous sampling methodologies (40,41). The use of quasi-experimental methods reduces confounding between IPV and long-term healthcare costs. These strengths and alignment of our findings with studies from other contexts and with our sensitivity analyses bolster the validity of our conclusions.

There are also several limitations. The HITS scale used to measure IPV, while having high sensitivity, specificity, and good concurrent validity with more comprehensive measures (47,79,80), lacks coverage of sexual violence, stalking, coercive control, and technology-facilitated violence. The extended HITS scale (79), which includes sexual IPV, was unavailable during the NEHW study. IPV exposure was only recorded at baseline, despite attempts to track later exposures using ICD-10 codes (T74.0-T74.9, X85-Y09, Z63.0). We hypothesize these codes are underutilized as they are not billable under OHIP (81). Given its historical underreporting, individuals in the comparison group may have experienced but not disclosed IPV (82,83); this may be mitigated since HITS behavior-based and avoids labeling IPV as either violence or abuse, which likely depresses reporting of IPV (84,85). Finally, costs were confined to OHIP data, omitting out-of-pocket or privately insured expenses, which are non-trivial components of healthcare spending in Canada.

## Conclusion

### Policy Implications

Our findings identify IPV as a significant burden on Canada’s healthcare system, emphasizing the need for policymakers to prioritize primary prevention as a cost-saving strategy. By addressing key social determinants (e.g., poverty, housing, gender inequality), governments with universal healthcare schemes can achieve long-term savings by reducing public healthcare costs for those at risk of IPV. This aligns with WHO and Canadian government recommendations for holistic, community-lead policies that prevent and mitigate IPV’s effects through social norms and healthcare improvements, rather than punitive actions for perpetrators (86,87).

Primary prevention approaches must also be paired with improving screening of, and service availability for, survivors (88), so they can be connected to trauma- and violence-informed legal, mental health, employment, and housing services (89,90). Access to transitional and long-term housing, in particular, has been cited as vital for women seeking to leave a violent relationship (91). For example, new models of care currently being tested by members of this research team aim to mitigate long-term health effects and costs by allowing women and children to remain in the family home, providing wraparound support in a “Safe at Home” model (92). Furthermore, since women experiencing IPV interact with the healthcare system more frequently, all healthcare professionals with direct patient contact should be trained to recognize signs of IPV and provide prompt and appropriate referrals and support through screening at mental health, emergency, and other types of healthcare system interactions in which IPV survivors are overrepresented. This should be substantiated with institutional and system-level policy changes that encourage the use of IPV-related ICD codes and allow them to be billed as the health outcomes they are.

Trauma-informed care and resource referral should be better integrated into healthcare systems, with policies at systems and institutional levels providing additional training and funding to wraparound services at discharge. Finally, additional IPV-related content in nursing, social work, and medical education enables professionals to improve patient-centered communication and care, potentially leading to earlier intervention in cases of violence (93).

### Future Directions

Women experiencing IPV use outpatient mental health services at higher rates.

Future studies could explore how IPV introduces delays and other barriers to mental healthcare. We also found evidence linking IPV with neighborhood-level material deprivation.

Future analyses could explore this relationship (using other neighborhood indicators) further, providing policymakers with evidence to develop effective community-based interventions that align and collaborate with healthcare systems.

### Conclusions

The healthcare costs measured in these analyses, while significant, are but a reflection of the psychological distress, physical pain, and disruption that survivors of IPV experience. The findings in this study therefore serve as a call to action for policymakers, community leaders, and healthcare providers to increase investment in research, primary prevention, and policy solutions that aim to prevent and mitigate IPV.

## Data Availability

All data produced in this present study is housed within the secure ICES Data & Analytic Virtual Environment (IDAVE). Access is only available with permission granted by ICES, formerly known as the Institute for Clinical Evaluative Science.

## Acknowledgements and Funding

Ethics approval was granted by the Research Ethics Board of [BLINDED FOR REVIEW] (REB# 20-120).

This work was supported by the Social Sciences and Humanities Research Council of Canada via an Insight grant to [BLINDED FOR REVIEW] (#435-2020-1410).

*Statement on the use of artificial intelligence (AI):* In the preparation of this manuscript, the AI tool ChatGPT, developed by OpenAI, was utilized for summarizing content, streamlining sentences, structuring sections, and answering specific queries related to the text. The use of ChatGPT facilitated the enhancement of the manuscript’s clarity and logical coherence. However, the authors assume full responsibility for the final content and ethical compliance with publication standards. ChatGPT was not used for the collection and analysis of data or the production of images or graphical elements.

