## Supplementals for "Long-term Public Healthcare Burden Associated with Intimate Partner Violence among Canadian Women: A Cohort Study"

### Appendix

#### Supplementary Tables & Figures

Supplementary Table 1: Evaluation of covariate Balance: standardized differences and variance ratios between treatment and control groups in the unmatched and matched samples for each model estimating average treatment effects (ATE) and average treatment effects on the treated (ATET) for cumulative cost and number of billings.

| | ATE: Log Cumulative Cost | | | | ATET: Log Cumulative Cost | | | | ATE: Cumulative Number of Billings | | | | ATET: Cumulative Number of Billings | | | |
|---|---|---|---|---|---|---|---|---|---|---|---|---|---|---|---|---|
| | **Standardized differences** | | **Variance ratio** | | **Standardized differences** | | **Variance ratio** | | **Standardized differences** | | **Variance ratio** | | **Standardized differences** | | **Variance ratio** | |
| | *Raw* | *Weighted* | *Raw* | *Weighted* | *Raw* | *Weighted* | *Raw* | *Weighted* | *Raw* | *Weighted* | *Raw* | *Weighted* | *Raw* | *Weighted* | *Raw* | *Weighted* |
| **Year of Birth** | | | | | | | | | | | | | | | | |
| | 0.3755 | 0.0213 | 1.0174 | 0.9549 | 0.3755 | -0.0160 | 1.0174 | 0.9379 | 0.3755 | 0.0214 | 1.0159 | 0.9539 | 0.3755 | -0.0162 | 1.0159 | 0.9374 |
| **Charlson Comorbidity Index** | | | | | | | | | | | | | | | | |
| | -0.2403 | -0.0242 | 0.7850 | 0.8875 | -0.2403 | 0.0250 | 0.7850 | 0.9559 | -0.2404 | -0.0243 | 0.7838 | 0.8863 | -0.2404 | 0.0252 | 0.7838 | 0.9553 |
| **Income** | | | | | | | | | | | | | | | | |
| <10,000 | 0.0240 | -0.0074 | 1.2971 | 0.9141 | 0.0240 | 0.0376 | 1.2971 | 1.5290 | 0.0239 | -0.0074 | 1.2952 | 0.9145 | 0.0239 | 0.0376 | 1.2952 | 1.5275 |
| 10,000-14,000 | 0.0668 | 0.0070 | 1.6098 | 1.0525 | 0.0668 | 0.0209 | 1.6098 | 1.1445 | 0.0666 | 0.0069 | 1.6075 | 1.0523 | 0.0666 | 0.0209 | 1.6075 | 1.1446 |
| 15,000-19,000 | 0.1492 | 0.0158 | 3.9886 | 1.1553 | 0.1492 | 0.0360 | 3.9886 | 1.2710 | 0.1491 | 0.0158 | 3.9828 | 1.1549 | 0.1491 | 0.0359 | 3.9828 | 1.2698 |
| 20,000-29,000 | -0.0586 | -0.0221 | 0.7774 | 0.9080 | -0.0586 | 0.0091 | 0.7774 | 1.0436 | -0.0590 | -0.0221 | 0.7763 | 0.9078 | -0.0590 | 0.0092 | 0.7763 | 1.0443 |
| 30,000-39,000 | 0.1134 | 0.0045 | 1.5295 | 1.0166 | 0.1134 | -0.0276 | 1.5295 | 0.9167 | 0.1130 | 0.0044 | 1.5273 | 1.0162 | 0.1130 | -0.0275 | 1.5273 | 0.9169 |

| | | | | | | | | | | | | | | | | |
|---|---|---|---|---|---|---|---|---|---|---|---|---|---|---|---|---|
| 40,000-49,000 | -0.0486 | -0.0056 | 0.8404 | 0.9797 | -0.0486 | 0.0321 | 0.8404 | 1.1355 | -0.0491 | -0.0056 | 0.8392 | 0.9796 | -0.0491 | 0.0321 | 0.8392 | 1.1355 |
| 50,000-74,000 | 0.0122 | -0.0044 | 1.0220 | 0.9924 | 0.0122 | 0.0015 | 1.0220 | 1.0025 | 0.0115 | -0.0043 | 1.0208 | 0.9926 | 0.0115 | 0.0015 | 1.0208 | 1.0026 |
| Unknown | -0.1336 | 0.0008 | 0.5581 | 1.0034 | -0.1336 | -0.0016 | 0.5581 | 0.9916 | -0.1340 | 0.0007 | 0.5573 | 1.0028 | -0.1340 | -0.0016 | 0.5573 | 0.9920 |
| **Neighborhood Material Deprivation** | | | | | | | | | | | | | | | | |
| | 0.1826 | 0.0011 | 1.2219 | 0.9109 | 0.1826 | -0.0020 | 1.2219 | 0.9042 | 0.1816 | 0.0011 | 1.2209 | 0.9116 | 0.1816 | -0.0020 | 1.2209 | 0.9048 |
| **Marital Status** | | | | | | | | | | | | | | | | |
| Unmarried | -0.0045 | -0.0198 | 0.9991 | 0.9910 | -0.0045 | 0.0318 | 0.9991 | 1.0144 | -0.0057 | -0.0199 | 0.9986 | 0.9910 | -0.0057 | 0.0320 | 0.9986 | 1.0145 |
| **Born in Canada**? | | | | | | | | | | | | | | | | |
| No | -0.1242 | 0.0077 | 0.9286 | 1.0042 | -0.1242 | -0.0242 | 0.9286 | 0.9828 | -0.1223 | 0.0075 | 0.9294 | 1.0041 | -0.1223 | -0.0244 | 0.9294 | 0.9827 |
| **Employment Status** | | | | | | | | | | | | | | | | |
| Unemployed | -0.0233 | 0.0074 | 0.9799 | 1.0070 | -0.0233 | 0.0478 | 0.9799 | 1.0488 | -0.0242 | 0.0074 | 0.9791 | 1.0070 | -0.0242 | 0.0479 | 0.9791 | 1.0490 |
| **Has Children** | | | | | | | | | | | | | | | | |
| No | -0.1595 | -0.0154 | 0.8896 | 0.9891 | -0.1595 | 0.0024 | 0.8896 | 1.0022 | -0.1606 | -0.0155 | 0.8891 | 0.9890 | -0.1606 | 0.0026 | 0.8891 | 1.0025 |

Note: Equal distribution between treatment and control groups is shown by standardized differences close to zero and variance ratios close to 1.

Supplementary Table 2: Estimated Average Treatment Effects (ATE) and Average Treatment Effects on the Treated (ATET) for outcomes over the study period (2011-2020) estimated with Propensity Score Matching. Exposure is IPV within 10-years of participation in NEHW.

| | Estimate | 95% Confidence Interval | |
|---|---|---|---|
| | Log of Total Healthcare Cost | | |
| **ATE** | 0.2042** | 0.0737 | 0.3347 |
| **ATET** | 0.1920* | 0.0445 | 0.3395 |
| | Total Number of OHIP Billings | | |
| **ATE** | 34.87* | 3.33 | 66.41 |
| **ATET** | 60.00*** | 26.54 | 93.45 |

*p-value < 0.05, **p-value < 0.01, ***p-value < 0.00
